# Feasibility, Acceptability and Potential Efficacy of the Group Problem Management Plus Intervention among adults in Kenya: A quasi-experimental study

**DOI:** 10.64898/2026.03.23.26349068

**Authors:** Patrick N. Mwangala, Kennedy Omondi, Amina Abubakar

## Abstract

**Aims:** The burden of common mental disorders (CMDs) is high in Kenya. Unfortunately, most Kenyans (75%) in need of mental healthcare cannot access these services. This study evaluates the feasibility, acceptability, and effectiveness of a brief, lay provider-delivered group-based psychological intervention – Group Problem Management Plus (gPM+) among adults with moderate symptoms of CMDs in a Kenyan urban informal settlement.

**Methods:** In this quasi-experimental pre-post study, 274 adults (63.5% females) in Changamwe sub-county in Mombasa were identified through screening of depressive symptoms (≥10 on the 9-Item Patient Health Questionnaire, PHQ-9), anxiety symptoms (≥10 on the 7-Item Generalized Anxiety Disorder Questionnaire, GAD-7) and symptoms of post-traumatic stress disorder - PTSD (≥3 on the Primary Care PTSD Screen for DSM-5, PC-PTSD-5). gPM+ comprised of 5 weekly group sessions with eight to twelve participants per group. The intervention was delivered by 10 trained non-specialist facilitators from a local civil society organization between August 2024 and April 2025. Primary outcomes were scores on PHQ-9, GAD-7 and PC-PTSD-5 assessed at baseline, 2 weeks, 5 weeks and 3 months follow-up. Secondary outcomes included functional impairment, self-identified problems, self-perceived social support, self-perceived wellbeing, and a measure of gPM+ acceptability, feasibility and appropriateness.

**Results:** 428 participants were screened for eligibility, of whom 274 (64%) participated in gPM+ at baseline and there were 241 (88.0%) participants at 3-months follow-up. The findings demonstrated that lay facilitators from a grassroots organization can be trained to achieve the desired competency to effectively implement gPM+ in an urban informal settlement under supervision. Overall, there was a good intervention uptake, with gPM+ considered appropriate and useful by participants and lay facilitators. Relative to baseline, the outcome evaluation indicated that at follow-up there was a statistically significant reduction in symptoms of depression anxiety and PTSD. We also observed statistically significant improvements in all secondary outcomes.

**Conclusion:** This formative study demonstrated robust acceptance of gPM+ in the community settings with delivery by lay facilitators under supervision. Preliminary evidence shows that gPM+ has the potential to improve the mental health and wellbeing of adults in urban informal settlements in Kenya. The study sets the stage for further exploration of the outcomes through large scale implementation and definitive randomised controlled trials in the community.

**Impact statement:** Most populations in sub-Saharan Africa (SSA) do not have adequate access to effective psychological and pharmacological interventions, with up to a 90% mental health treatment gap reported. Evidence-informed psychological interventions have been found effective in some SSA countries. Nonetheless, there are significant barriers to the sustainable delivery of such interventions in the region, including limited mental health funding and infrastructure, inadequate psychological treatments adapted to the local context, and challenges associated with their implementation, e.g., limited availability of mental health specialists. To bridge this gap in Kenya, the current study evaluates the feasibility, acceptability, and potential efficacy of a brief, lay provider-delivered group-based psychological intervention – Group Problem Management Plus (gPM+) among adults with moderate symptoms of CMDs in a Kenyan urban informal settlement. This approach holds promise in scaling up access to mental health services in low-resource settings, e.g., through integration with local grassroots organizations, task-shifting, and leveraging local solutions/expertise.

## Introduction

Since the first Global Burden of Disease Study (GBD) findings were published in 1990 (Murray 2022; Murray et al. 1994), there has been increasing evidence that: (a) mental disorders constitute a leading cause of disease-related burden worldwide, (b) a major part of this burden (∼70%) is concentrated in low and middle-income countries (LMICs), (c) there has been no evidence of a reduction in the global prevalence or burden since 1990 despite compelling evidence of interventions (Collaborators GMD 2022). Within sub-Saharan Africa, the burden of mental and substance use problems is estimated to increase by 130% by 2050, partly due to population growth and ageing (Charlson et al. 2014). Despite the high burden of mental health problems, and the adverse impact of these disorders, mental health infrastructure, especially in LMICs, is not adequately resourced to meet the current and growing demand for care. Most mental healthcare services are concentrated at the apex of the service paradigm (referral hospitals and specialist services), with fewer resources available at the community level. As such, over 75% of individuals living with mental health problems in many parts of SSA, including Kenya, do not receive treatment, and those who receive treatment are not always retained in care, and experience variable access to quality services.

To address the limited provision of mental health services in LMICs, mental health experts and the World Health Organization (WHO), have developed brief, scalable transdiagnostic low-intensity psychosocial interventions within its Mental Health Gap Action Programme (World Health Organization 2008). One such program is Problem Management Plus (PM+), that was introduced in 2015 and aims to alleviate symptoms of common mental health problems among adults (Dawson et al. 2015). Being a task-shifted approach, PM+ can be delivered by trained and supervised non-specialist facilitators (also called helpers). Since its introduction in 2015, PM+ has been adapted and implemented in about 20 countries across the world and shown to be acceptable, feasible, and effective in improving symptoms of CMDs in both individual and group formats in communities affected by adversity (Cai et al. 2025; MwangalaMakandi et al. 2024; Schaefer et al. 2023; Zhang et al. 2025). The group format of PM+ (gPM+) has the potential to reach a higher number of people and therefore is more cost-effective for LMICs. A recent meta-analysis of gPM+ (Zhang et al. 2025) demonstrated that gPM+ improved physical functioning, and reduced negative emotions. Nonetheless, the authors found no statistically significant improvements in social-interpersonal level or reduction of antecedent and post-traumatic stress disorders – and thus recommended further studies to verify the efficacy of gPM+ for people experiencing psychological distress.

Like many countries in SSA, Kenya’s public healthcare sector currently offers inadequate mental health services (Bitta et al. 2017; Kwobah et al. 2023). Prevalence estimates show that about 25% and 40% of outpatients and inpatients suffer from mental health problems respectively (Ministry of Health 2024). Higher prevalence estimates have been reported among youth (Mbithi et al. 2023) and women (Mwangala et al. 2025) residing in urban informal settlements. Although Kenya has a comprehensive primary healthcare system (Ministry of Health 2020a), mental healthcare services are largely non-existent within the primary care. A recent report on the state of mental healthcare in the country (Ministry of Health 2020b) showed that 27 out of the 47 counties in the country do not have a dedicated mental/psychiatric units. Besides, the few mental health providers are concentrated in a few psychiatric institutions and high-level hospitals, which tend to be overcrowded and largely focus on severe mental disorders, highlighting the huge gap of community mental health services and an optimal referral pathway. The government has embraced the contributions of collaborative care models such as task shifting by instituting the national framework for the implementation of PM+ in the country (Ministry of Health 2018). So far, two studies have evaluated the feasibility and effectiveness of individual-based PM+ in Kenya (Bryant et al. 2017; Nyongesa et al. 2022). The first one evaluated the effectiveness of PM+ on psychological distress among women with a history of gender-based violence in urban Kenya (Bryant et al. 2017), while the second study adapted the individual-based PM+ for telephone delivery and preliminarily evaluated its acceptability and feasibility among young people living with HIV (Nyongesa et al. 2022). To the best of our knowledge, no study in Kenya has formally evaluated the feasibility and effectiveness of the gPM+.

To address the significant mental health needs of adults residing in Kenya, and particularly women residing in urban informal settlements, we conducted a quasi-experimental pre-post study of gPM+ to adults living in an urban informal settlement at the coast of Kenya. The aim of this study was to evaluate the feasibility, acceptability and potential efficacy of gPM+ among adults with moderate symptoms of CMDs. A key feature of this study was to explore the feasibility of delivering this intervention by trained and supervised lay facilitators drawn from staff/representatives of a local grassroots organization in the community.

## Methods

### Study design and setting

This was a quasi-experimental pre-post study conducted between April 2024 and April 2025 in Portreitz, Chaani and Kipevu wards – all located within Changamwe sub-county at the coast of Kenya. The study was conducted in partnership with Mizizi Youth Organization, a community-based mental health organization located in Mombasa County, who were the local implementers of gPM+ within Changamwe subcounty. Mombasa county is part of the 47 devolved units of government in Kenya which came into effect in 2013. Changamwe sub-county is one of the six sub-counties in Mombasa County. According to the most recent population and housing census of 2019, Changamwe sub-county had a population of 131,882 people (Kenya National Bureau of Statistics 2019). A significant proportion of these residents (about 40%) live in informal settlements with limited access to water, insecure tenure, inadequate housing, poor environmental conditions and high crime rates. The project was prospectively registered (Pan African Clinical Trial Registry, no. PACTR202406464406701).

### Participants

Participants comprised of adult men and women resident in Changamwe sub-county at the time of recruitment which took place between August 2024 and November 2024. Potential participants were recruited by the implementing partner, Mizizi youth organization. Mizizi is a grassroots organization that has been in operation in Changamwe since 2020, providing community mental health services (largely promotive and low-intensity psychological interventions) to adolescents, young people and other members of the public. Before recruitment, staff of Mizizi youth organization held a consultative stakeholder meeting at their office in Changamwe. The meeting brought together key community representatives including local administrators (chiefs), village elders, and youth champions from different grassroots organizations and introduced the gPM+ to the community to facilitate community buy-in and establish collaborative frameworks for successful participant recruitment and successful project implementation. Following the stakeholders meeting, 10 Mizizi representatives carried out extensive community engagement sessions with different community members including local leaders, healthcare providers and religious leaders to ensure community awareness and their participation in the program. Sensitization activities included talks in public barazas and community outreach programs. Mobilization efforts were carried out with the help of village elders, gPM+ facilitators and youth champions drawn from various grassroots organizations drawn from the three wards.

Participants were identified through household visits, workplaces, community referrals, local health centers, and outreach programs. Potential participants were identified and invited to participate in the screening process if they meet the following criteria: (a) aged ≥ 18 years old, (b) residence in Changamwe Sub-county, (c) scores of ≥10 on the 9-Item Patient Health Questionnaire (PHQ9) (Kroenke et al. 2001), (d) scores of ≥10 on the 7-Item Generalized Anxiety Disorder Scale (GAD7) (Spitzer et al. 2006), and (e) scores of ≥3 on the Primary Care Post Traumatic Stress Disorder screen for DSM-5 (PC-PTSD-5) (Prins et al. 2016). PHQ9, GAD7 and PC-PTSD-5 assess symptoms of depression, generalized anxiety, and post-traumatic stress respectively. Exclusion criteria consisted of having (i) any acute medical condition, (ii) pending risk of suicide (gPM+ manual suicidality assessment), (iii) signs and symptoms of a severe mental disorder such as psychotic disorders or substance use dependence, (iv) severe neurocognitive impairment (gPM+ manual checklist). Excluded participants were referred to access mental health services through the sub-county mental health referral pathway.

Participants’ informed consent involved two steps: (a) consent to participate in the screening process, and (b) participants who screened positive were then invited to provide their consent to participate in the gPM+ intervention. All respondents completed a written consent form, and those who were illiterate provided witnessed oral consent in line with WHO recommendations (Bhutta 2004).

### Procedures

The individual version of PM+ intervention has previously been adapted and implemented in Kenya among young people at the coast of Kenya (Nyongesa et al. 2022), and women with a history of gender-based violence in Nairobi (Bryant et al. 2017). To the best of our knowledge, the gPM+ has not been formally assessed in Kenya. As such, prior to the implementation of the current study, the translation and cultural adaptation of gPM+ was reviewed in a workshop with a PM+ expert, translators, menta health researchers and representatives from civil society organizations (CSOs) to ensure that the assessment tools and intervention were appropriate for the study setting and population.

The group version of PM+ is an adaptation of the WHO individual PM+ (Dawson et al. 2015). gPM+ consists of 5 weekly 2-hour group sessions delivered by non-specialist helpers. In each session, participants are taught four strategies. The first session incorporates psychoeducation and stress management through a slow breathing exercise, session two focusses on learning a structured problem management strategy, session three is on behavioral activation and session four is on strengthening social support. The fifth and last session reviews the four strategies and addresses relapse prevention. The focus of group sessions is to teach participants skills in managing stressors in the community and families, and group discussion is facilitated to encourage members to share their experiences on problem-solving. In the current project, gPM+ sessions were usually conducted in different community spaces including schools, churches and youth friendly centers. In some instances, a few of the sessions took place in the participants’ homes. gPM+ groups comprised of 8-12 participants, separated by gender and each session was facilitated by two gPM+ helpers.

Ten representatives (6 female) from Mizizi Youth Organization were engaged to implement gPM+. These facilitators had at least 12 years’ school education and six months’ experience in community mental healthcare e.g. psychoeducation, and other forms of mental health promotion. These facilitators were provided with a 96-hour training programme (delivered by a PM+ master trainer) over two weeks. Three local supervisors who were experienced mental health practitioners were also trained in gPM+. Training covered different topics including knowledge on common mental health problems, basic helping skills, ethics, gPM+ intervention protocol, group facilitation skills, selfcare strategies, psychological first aid, suicide risk assessment and response, and local referral pathways. As part of the gPM+ training, facilitators completed three practice cycles, as lead facilitator and co-facilitator under close supervision. Competency was evaluated using mock interviews that assessed key strategies e.g. verbal and non-verbal communication, rapport, and clarity of gPM+ strategies.

During the project, facilitators received 1.5 hours of weekly (at first then biweekly) supervision by a local supervisor virtually. Supervision included a review of clients’ progress and individual case management, refresher training on gPM+ strategies and enhancing facilitators’ wellbeing. Intervention fidelity was monitored by an independent observer in 10% of randomly selected sessions against a standardized checklist comprising gPM+ strategies. Based on this evaluation, each strategy was evaluated as present/absent, and whether it was provided satisfactorily or not. Weak areas that were identified were reinforced during the supervisions.

### Outcome measures

We assessed three primary outcomes in this project that included a) anxiety symptom score, measured with GAD7 (Spitzer et al. 2006), b) depressive symptom score, measured with PHQ9 (Kroenke et al. 2001), and c) PTSD symptom score, measured with PC-PTSD-5 (Prins et al. 2016). All the three measures were administered at baseline, 2-week post intervention, 5-week post intervention and 3-months follow-up. Higher scores indicate more anxiety, depression and PTSD symptoms than lower scores. The three measures have been validated across different cultures, including Kenya and have been shown to have good reliability and validity (MwangalaGuni et al. 2024; Mwangi et al. 2020; Nyongesa et al. 2020).

Secondary outcomes included perceived social support, general wellbeing, functional impairment, changes in self-identified problems for which the person sought help, and gPM+ measures of acceptability, appropriateness and feasibility. Apart from the latter which was measured at the 5-week endpoint, the rest of the secondary outcomes were assessed at baseline, 2-week postintervention, 5-week endpoint and 3-months follow-up. Perceived social support was assessed using the 12-item Multidimensional Scale of Perceived Social Support – MSPSS (Zimet et al. 1988). General wellbeing was measured with the World Health Organization 5-item well-being index – WHO5 (Chongwo et al. 2018) while functional impairment was measured using the 12-item World Health Organization Disability Assessment Schedule (Üstün et al. 2010). The Psychological Outcome Profiles (PSYCHLOPS) was used to measure changes in problems for which the person sought help (Ashworth et al. 2005). gPM+ acceptability, appropriateness and feasibility were measured using the Acceptability of Intervention Measure (AIM), Intervention Appropriateness Measure (IAM), and Feasibility Intervention Measure (FIM) (Weiner et al. 2017) respectively. Additionally, we administered a 9-item exit checklist documenting the participants’ experiences with gPM+. We also conducted one focus group discussion with the lay facilitators at the end of the intervention to explore their experiences delivering gPM+. All outcome assessments were administered by the trained gPM+ facilitators. Information on adverse events e.g. suicide attempts, admissions and violence were also documented.

### Statistical analysis

This project was primarily formative, with the intention of assessing the acceptability, appropriateness, feasibility and potential efficacy of the gPM+ in Kenya as delivered by trained non-specialist facilitators from the local community. Thus, no formal sample size and power calculations were carried out (Leon et al. 2011). Feasibility assessment of key components, e.g. training, delivery and supervision, requires the non-specialist facilitators to have a sufficient caseload. The chosen sample size allowed each facilitator to have a minimum of 3 gPM+ sessions in a week. Given the initial evidence of PM+ effectiveness in a similar setting in Kenya (albeit with the individual format) (Bryant et al. 2017), high burden of common mental health problems in this setting (Mwangala et al. 2025), and the huge mental health treatment gap in Kenya as a whole (Ministry of Health 2020b), we included a relatively large sample of participants in this feasibility study.

Data analysis was carried out in STATA version 17 for Windows (StataCorp LP, College Station, Texas, USA). Descriptive statistics e.g. frequencies, percentages, and means (standard deviations) were utilized to summarize the sample characteristics, as well as the acceptability, appropriateness and feasibility of gPM+. Qualitative data exploring the experiences of delivering gPM+ by lay facilitators was analyzed using thematic analysis. We used paired sample t-tests to evaluate the potential effect of the gPM+. We also derived Cohen’s d effect size by calculating the mean difference between assessments and dividing this by the pooled standard deviation.

## Results

### Participants characteristics

Participants were recruited to the study between August and November 2024. Follow-p assessments were completed in April 2025. A total of 428 participants were screened, of whom 274 were found to be eligible and consented to take part in the gPM+. Out of 428, 119 were excluded because of not meeting the inclusion criteria, 33 declined participation and 2 participants presented with suicidal ideations and were excluded. The study flowchart is given in **Figure 1. Table 1** highlights the sociodemographic characteristics of the study participants at baseline. The median age of the respondents was 27 years (IQR 22 – 37). Majority of the participants were female (64%), had basic education (95%), were Christians (69%), unemployed (67%), with debts (75%), and experiencing food insecurity (84%).

**Table 1.** Baseline characteristics of the study participants, *n =* 274.

| **Variable** | ***n* (%) or median (IQR)** |
| --- | --- |
| Median age, years | 27 (22 – 37) |
| Female sex *n* (%) | 174 (63.5%) |
| **Educational level** |  |
| *None* | 3 (1.1%) |
| *Primary level* | 77 (28.1%) |
| *Secondary level* | 179 (65.3%) |
| *Tertiary level* | 15 (5.5%) |
| **Religion** |  |
| *Christian* | 189 (69.0%) |
| *Muslim* | 85 (31.0%) |
| **Marital status; OM=15** |  |
| *Never married* | 118 (43.1%) |
| *Married/cohabiting* | 108 (39.4%) |
| *Separated/divorced* | 28 (10.2%) |
| *Widowed* | 5 (1.8%) |
| **Employment** |  |
| *Skilled/professional* | 5 (1.8%) |
| *Unskilled/casual* | 85 (31.0%) |
| *Unemployed* | 184 (67.2%) |
| **Monthly household income (Ksh)** |  |
| <10000 | 246 (89.8%) |
| 10000 – 29999 | 23 (8.4%) |
| >30000 | 5 (1.8%) |
| Household debt (yes) | 206 (75.2%) |
| Household food insecurity in the past 3 months (yes) | 231 (84.3%) |
| **Living arrangements; OM=9** |  |
| *Own house (not paying rent)* | 15 (5.5%) |
| *In a rented house* | 250 (91.2%) |
| *OM = Observations with missing values* |  |

**Figure 1.**
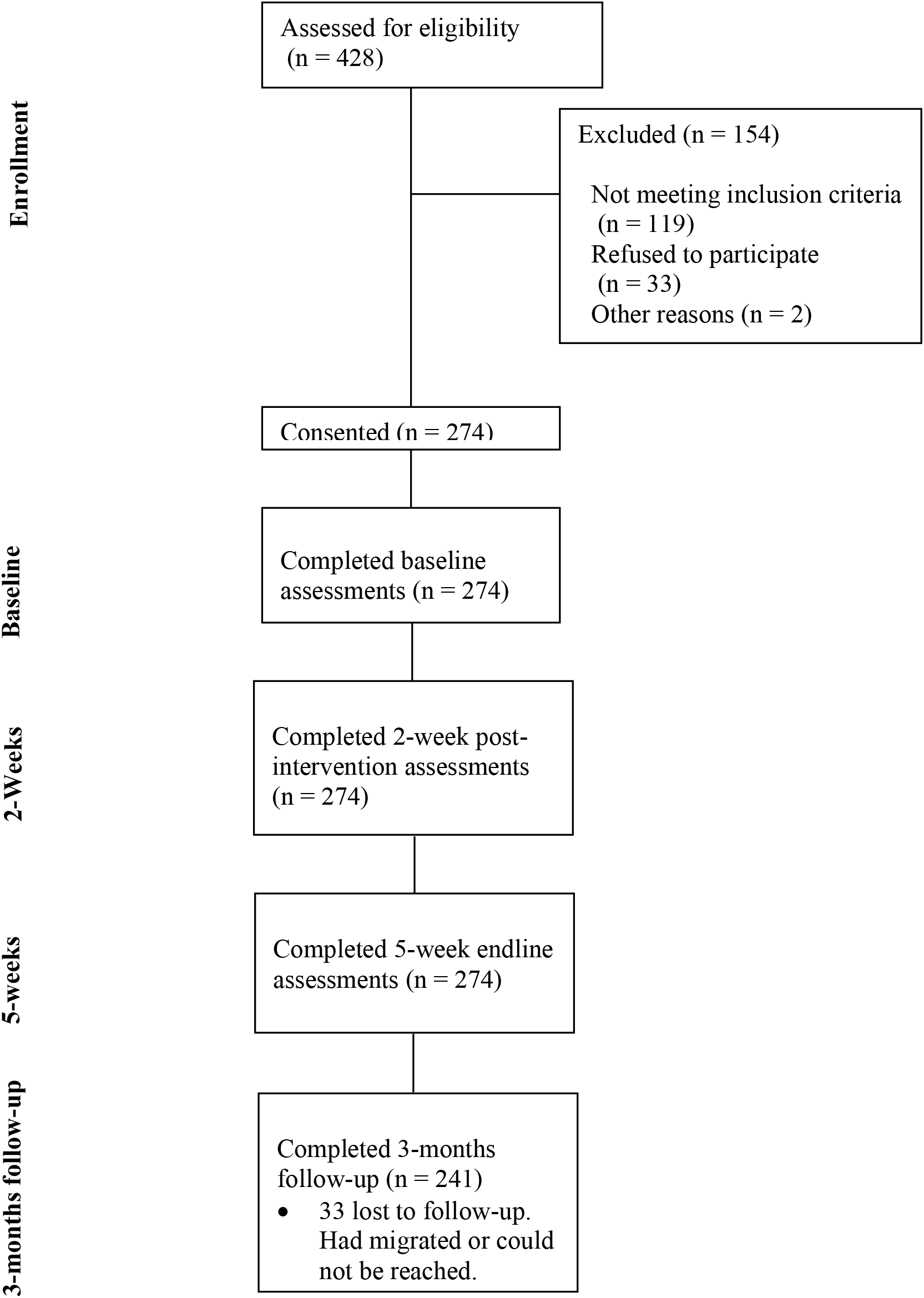
Participants flow diagram

### Attendance, follow up and fidelity

All the participants who began the intervention at baseline completed the full five gPM+ sessions. However, at the 3 months follow up, 33 (12%) of the participants were lost to follow up. Most of these participants could not be reached on phone while others had migrated or were busy. As indicated above, two participants were excluded at the screening phase due to suicidal ideation; both were referred to specialized services for appropriate treatment. No adverse events were reported throughout the study. 10% of the sessions were assessed using the gPM+ fidelity checklist and indicated that 91% of the core components of gPM+ were delivered well and adhered to.

### Acceptability, Appropriateness and Feasibility of the gPM+, and User satisfaction

Measures of acceptability, appropriateness and feasibility were administered to all participants at the 5-week endline assessment. Participants rated their responses on five-point Likert scales, with 5 being the highest (with total scores ranging from 0 to 20). Clients rated gPM+ as acceptable (mean 18.7, SD=2.0) appropriate (mean 18.9, SD=1.5) and feasible (mean18.8, SD=1.6). Regarding user satisfaction, we administered a dichotomous (yes/no) exit interview schedule to all participants at the 5-week endline assessment. All participants (100%) reported to have enjoyed the training, 99% felt satisfied after the intervention, and all the participants felt the gPM+ strategies were easy to understand. A further 99% of the participants found the intervention to be therapeutic in their personal lives, and the sessions fit within their weekly schedule easily and all of them expressed willingness to use gPM+ strategies outside the study e.g. future personal challenges and helping others.

### Experience of delivering gPM+

Generally, the facilitators shared positive experiences of the gPM+ intervention regarding their interaction with the intervention participants, their roles as delivery agents and likely effects of the intervention in their own lives. They all reported that they were able to develop and sustain therapeutic relationships with their clients. However, they noted that it was somehow difficult for clients to open up at first, but assurances of confidentiality of information, and shared experiences with the other participants encouraged them to open up. The facilitators also identified positive changes gPM+ had made for them personally e.g. in capacity building which made them knowledgeable about effects of common mental disorders and better equipped to serve their communities and mitigate their own problems. They were all clear on the potential value of scaling up gPM+ in the community and the positive impact it could have on the population. As a grassroot organization, they were keen on integrating this programme as part of their work packages in the community. Some of the facilitators indicated that they had been approached by community members and leaders that had heard about gPM+ and wanted to participate or mentioned others who could benefit from the programme, suggesting gPM+ was positively received by the community.

### Evaluation of primary and secondary outcomes

Table 2. highlights the mean primary and secondary outcomes (standard deviation) at each observation time point across the study.

#### Primary outcomes

We observed a consistent significant reduction of depressive, anxiety and PTSD symptoms across all the observation time points. The mean change in depressive symptoms from baseline to 3 months follow-up was 9.2 [95% CI, 8.6 to 9.7], *p =* <0.001. Similar patterns were observed for anxiety symptoms (mean difference 7.3 [95% CI, 6.7 to 7.8], *p =* <0.001) and PTSD symptoms (mean difference 2.6 [95% CI, 2.3 to 2.9], *p =* <0.001). These results indicated large effect sizes for depressive symptoms (1.3 [95% CI, 1.1 to 1.5]), anxiety symptoms (1.2 [95% CI, 1.0 to 1.4]) and PTSD symptoms (2.1 [95% CI, 1.8 to 2.3]) as measured by Cohen’s d.

**Table 2.**
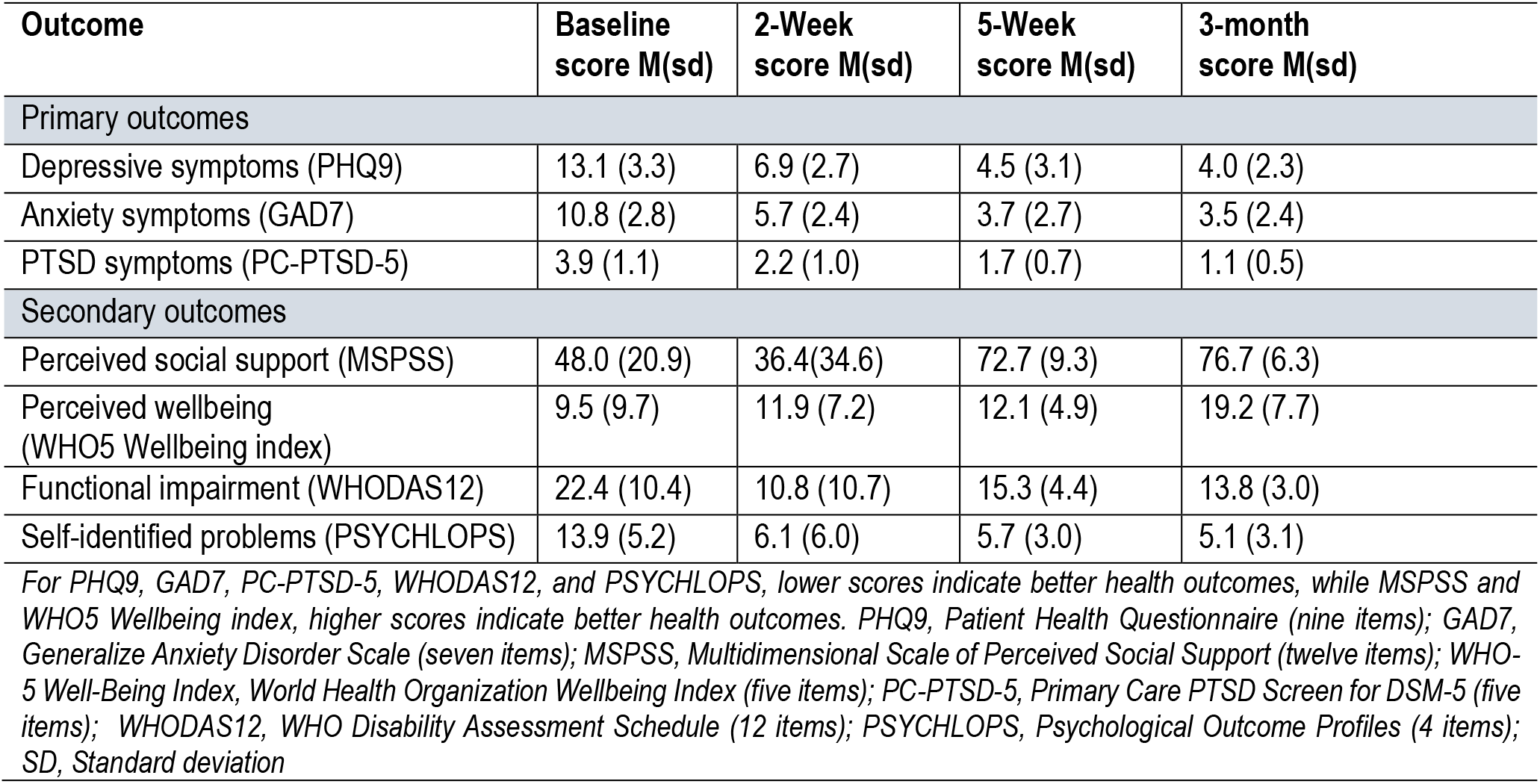
Mean primary and secondary outcomes at baseline, and post-intervention across the study.

| Outcome | Baseline score M(sd) | 2-Week score M(sd) | 5-Week score M(sd) | 3-month score M(sd) |
| --- | --- | --- | --- | --- |
| Primary outcomes |  |  |  |  |
| Depressive symptoms (PHQ9) | 13.1 (3.3) | 6.9 (2.7) | 4.5 (3.1) | 4.0 (2.3) |
| Anxiety symptoms (GAD7) | 10.8 (2.8) | 5.7 (2.4) | 3.7 (2.7) | 3.5 (2.4) |
| PTSD symptoms (PC-PTSD-5) | 3.9 (1.1) | 2.2 (1.0) | 1.7 (0.7) | 1.1 (0.5) |
| Secondary outcomes |  |  |  |  |
| Perceived social support (MSPSS) | 48.0 (20.9) | 36.4(34.6) | 72.7 (9.3) | 76.7 (6.3) |
| Perceived wellbeing (WHO5 Wellbeing index) | 9.5 (9.7) | 11.9 (7.2) | 12.1 (4.9) | 19.2 (7.7) |
| Functional impairment (WHODAS12) | 22.4 (10.4) | 10.8 (10.7) | 15.3 (4.4) | 13.8 (3.0) |
| Self-identified problems (PSYCHLOPS) | 13.9 (5.2) | 6.1 (6.0) | 5.7 (3.0) | 5.1 (3.1) |
| For PHQ9, GAD7, PC-PTSD-5, WHODAS12, and PSYCHLOPS, lower scores indicate better health outcomes, while MSPSS and WHO5 Wellbeing index, higher scores indicate better health outcomes. PHQ9, Patient Health Questionnaire (nine items); GAD7, Generalize Anxiety Disorder Scale (seven items); MSPSS, Multidimensional Scale of Perceived Social Support (twelve items); WHO-5 Well-Being Index, World Health Organization Wellbeing Index (five items); PC-PTSD-5, Primary Care PTSD Screen for DSM-5 (five items); WHODAS12, WHO Disability Assessment Schedule (12 items); PSYCHLOPS, Psychological Outcome Profiles (4 items); SD, Standard deviation |  |  |  |  |

#### Secondary outcomes

Similar patterns of improvement were observed for all secondary outcomes at 3 months (**Table 2**). The mean change (SD) in perceived social support scores from baseline to 3 months follow-up was -24.3 (18.0) while that of general wellbeing was -8.7 (8.6). On the other hand, the mean change (SD) in functional impairment scores from baseline to 3 months follow-up was 10.0 (8.3) while that of self-identified problems was 10.2 (4.5). All the differences were statistically significant (*p*<0.001) and exhibited medium to large effect sizes.

## Discussion

### Summary of key findings

This study assessed the acceptability, appropriateness, feasibility and initial efficacy of the gPM+ intervention, delivered by non-specialists to adults with moderate levels of common mental health problems in an urban informal settlement in Kenya. The findings show that lay facilitators from a local grassroot organization can be trained to achieve the desired competency to effectively implement the intervention in the community under supervision. Overall, intervention uptake was excellent, with gPM+ considered appropriate, acceptable and therapeutic by participants and lay facilitators. The outcome evaluation, which was not powered, nonetheless demonstrated relevant improvements in symptoms of anxiety, depression, PTSD, functioning, social support, general wellbeing and self-identified problems across all the timepoints. The feasibility of assessments, procedures and intervention suggests that a fully powered gPM+ evaluation trial is achievable in this context.

### Comparison with findings from other studies

To the best of our knowledge, this is the first formative study in Kenya to look at the appropriateness, acceptability, feasibility and potential efficacy of the gPM+. Previous PM+ studies in this setting have examined the acceptability, feasibility and effectiveness of the individual version of PM+ among young people at the coast of Kenya (Nyongesa et al. 2022) and women with a history of gender-based violence in urban Kenya (Bryant et al. 2017; Van’t Hof et al. 2018). Our findings add to the body of research evidence on PM+ in Kenya and further supports the implementation and dissemination of this low-intensity psychological intervention among adults facing adversities in urban informal settlements. Notably, group-based programs may be more cost-effective than individually implemented mental health interventions (Huntley et al. 2012), and when they can be successfully delivered by lay facilitators, this further enhances the likelihood that programs like gPM+ could be implemented in informal settlements in low-resource countries (Hamdani et al. 2020).

In LMICs, the use of trained and supervised lay counsellors for the implementation of mental health interventions has been preferred given the high treatment gap and severe shortage of mental health specialists (Chibanda et al. 2011; Van Ginneken et al. 2011). This study extends the growing evidence in support of task-shifting approaches in the delivery of evidence-based psychological interventions (Van Ginneken et al. 2011). The high acceptability and successful implementation of this feasibility study suggests that lay facilitators can effectively deliver gPM+ after training and continued supervision. Local religious and administrative leaders introduced the lay facilitators to the community as part of community engagement before participants’ recruitment began. This was ideal as they are trusted and respected in the communities, overcoming a likely barrier in accessing study participants. Additionally, our partnership with representatives of a grassroot organization as implementers of gPM+ likely ensured acceptability and an ability to relate well to their participants. There is evidence showing participants can have preference for delivery agents from within their localities over outside agents, as they are easily accessible and reduce the stigma attached to help seeking (Singla et al. 2014). Moreover, local selection is considered to increase the potential for future intervention scale-up. The lay facilitators had at least 12 years of education, and with some experience in community mental health promotion, enabling access to a large pool of potential people who can take up this role. Furthermore, locally selected facilitators could be more conscious of the local context, which is important in intervention implementation. The use of lay facilitators was also found to be acceptable in previous PM+ exploratory studies in Kenya (Nyongesa et al. 2022; Van’t Hof et al. 2018).

The supervision models utilized in this study are potentially scalable, with a few specialists in another location able to provide group supervision to several facilitators through virtual meetings. Moreover, the group format of supervision enabled lay facilitator support and supervision, which was found to be essential in managing difficult situations during field activities e.g. facilitators’ psychological wellbeing.

The high recruitment of respondents into the project supports the feasibility of enrolling adults in urban informal settlements into gPM+ intervention. We observed a 100% retention at the end of the intervention, and a good retention of 88% at 3-months follow-up, which is higher than that reported in previous formative studies (Nyongesa et al. 2022). It is important to note however, that the recruitment of male participants was met with hesitancy at first due to their belief that only those with severe mental illnesses needed support, their busy schedules and fear of sharing their issues with others, especially the opposite gender. Similar observations have been reported elsewhere and emphasize the importance of using de-stigmatizing local idioms and language during the planning and community engagement phase with different stakeholders (Kohrt and Hruschka 2010). As experienced by the facilitators in this project, sensitization meetings, worked to normalize experiencing adversity and distress, as well as to emphasize that gPM+ was for those experiencing common mental health problems. To enhance the recruitment of men, facilitators expanded their recruitment places to include workplaces, and areas mostly frequented by men. During the implementation of the programme, we held separate groups for men and women. The overall feasibility and acceptability of conducting the intervention amongst men indicates that it is possible to include both genders in a large trial in the Kenyan context, with some potential barriers in recruitment.

Exit interviews with participants revealed that all gPM+ strategies were contextually appropriate. This was corroborated by the qualitative interviews with the lay facilitators. When implemented, we observed a consistent trend towards a reduction in symptoms of common mental disorders across all timepoints, which was statistically significant relative to baseline assessments although the study was not powered to detect clinically significant differences on outcome measures. We also observed a similar pattern for the other psychosocial and functional outcomes, i.e. social support, functional impairment, general wellbeing and self-identified problems. These results are consistent with previous findings in Kenya (Dawson et al. 2016; Nyongesa et al. 2022; Van’t Hof et al. 2018) and other settings (Acarturk et al. 2022; Sangraula et al. 2020).

### Interpretation of the current findings and implications for future work

The results of this study suggest that the implemented gPM+ appears contextually appropriate in an urban informal settlement at the coast of Kenya. Additionally, the use of lay facilitators drawn from a local community-based organization is also feasible and acceptable. Additionally, it is feasible to recruit and retain participants in the gPM+ intervention, evidenced by the good response rate and retention rates, with some potential barriers in the recruitment of men. It is also feasible to implement five weekly sessions of about 2 hours each via face-to-face meetings. Initial feasibility data indicates that gPM+ has the potential to alleviate symptoms of common mental disorders and improve participants’ psychosocial and functional wellbeing. The findings from this study give preliminary evidence important in guiding the community implementation of gPM+ in this setting. There may be need to assess and ascertain to what extent lay facilitators can retain the gPM+ knowledge and skills for continued therapy in the community. A further qualitative evaluation of gPM+ with different stakeholders, e.g. local leaders, gPM+ participants and providers, county ministry of health officials, and healthcare providers, can provide additional insights into the possible barriers and facilitators of implementing gPM+ in this setting.

### Limitations

We acknowledge some limitations that should be considered when interpreting the results of this study. Although we observed significant differences at different time points due to gPM+ intervention, sample size and power calculations were not carried out to detect such differences during the design of this study, as this was primarily formative work. Relatedly, the pre/post design carries with it inherent challenges compared to other experimental designs. It is not possible to fully attribute the improvements in mental health and wellbeing outcomes over the course of this intervention to the intervention itself. However, the timeline of the assessments – make other potential explanations for the observed improvements quite unlikely.

## Conclusions

Our findings in this study indicate that the methodological procedures, including community engagement, participants recruitment, retention and attendance to gPM+ sessions, and the training and supervision of lay facilitators are promising and can guide a fully powered RCT in this population. Taken together, gPM+ seems to be appropriate, and acceptable to adults facing psychosocial vulnerabilities in this urban informal settlement, and feasible for lay facilitators to deliver the intervention. This evidence adds to the body of research in support of task-shifting approaches in delivering evidence-based psychological interventions and provides an important foundation for the scale-up of mental health services in this setting in addressing the huge mental health treatment gap in LMICs.

## Data Availability

All data produced in the present study are available upon reasonable request to the authors

## Data availability statement

The de-identified dataset used and analyzed during this current study will be made available upon reasonable request, in due consideration of Aga Khan University’s data sharing policies. Requests to access the datasets should be directed to the Research Office Aga Khan University Kenya through.

## Acknowledgements

The authors would like to thank all the stakeholders and participants who contributed to this study. Special appreciation goes to Rachael Odhiambo and the representatives of the Mizizi Youth Organization who implemented the intervention: Ramadhan Omar, Rachel Muthoni, Irene Okwado, Geoffrey Arasa, Kennedy Omondi, Elizabeth Mwongeli, Dama Kahindi, Collins Owuor, Florence Mghenyi, and Rachel Ngado. We would like to appreciate the Mombasa County Government – department of health services officials for their invaluable support in providing local expertise.

## Author contribution

AA conceptualized and obtained funding for the study. AA and PNM developed study protocols, tools, and training programmes. PNM, and KO coordinated the data collection process. PNM wrote the first draft of the manuscript. All authors critically reviewed subsequent versions of the manuscript and approved the final version for submission. All authors read and approved of the final manuscript.

## Ethics statement

The project was carried out in accordance with ethical principles and guidelines for involving human participants as stipulated in the Helsinki Declaration. Ethical approval was sought and given by the Aga Khan, Nairobi Institutional Scientific and Ethics Review Committee (Ref: 2023/ISERC-37(v2)). Permission to conduct the study in Kenya was granted by the National Commission for Science, Technology and Innovation (Ref: NACOSTI/P/23/28548). All the participants provided written informed consent for their participation in the study.

